# GENETIC COUNSELING IN THE MIDDLE EAST

**DOI:** 10.1101/2023.09.07.23292346

**Authors:** Shruti Shenbagam, Alan Taylor, Ruchi Jain, Khalid A. Fakhro, Fowzan S. Alkuraya, Ahmad N. Abou Tayoun

## Abstract

Genomic advancements have led to increased utilization of genetic testing in clinical care, yet barriers to accessing genetic counseling and genomics services remain, particularly in the Middle East where inherited diseases are highly prevalent due to consanguinity. Limited knowledge of healthcare professionals’ experiences in genetic counseling in the Middle East necessitates understanding their perspectives for better service improvement in the region.

A survey of 32 healthcare professionals providing genetic counseling services in the Middle East explored provider experiences, patient attitudes and cultural/psychosocial factors related to genetic testing. Among the respondents, 21 providers (65.6%), caring for patients of multiple ethnicities, including Arabs, recognised that there are unique challenges to counseling between these patient groups. Thematic data analysis identified that higher levels of consanguinity and stoic nature of the people are unique cultural considerations for this region. Language barriers and limited resources were identified as genetic counseling challenges. Overall, patients in the region demonstrated good coping abilities with a genetic diagnosis. Eighteen responses (56%) highlighted an overall positive attitude, with increasing awareness and acceptance towards genetic testing in this region. This study highlights the need for further research and interventions to address the unique challenges and improve genetic counseling services in the Middle East.

**What is known about this topic?:** Genetic counseling is typically provided by geneticists and genetic counselors, but due to the limited numbers in the Middle East, this service is also provided by other physicians across various specialties. There is limited knowledge regarding the experiences of healthcare providers offering genetic counseling services in the Middle East and the specific challenges they face in this region.

**What this paper adds to the topic?:** Our study offers valuable insights into the perspectives of healthcare providers regarding patient attitudes, cultural considerations, and psychosocial factors in genetic testing, with the aim of improving genetic counseling services and enhancing patient outcomes. These findings, along with our recommended strategies shed light on the unique dynamics of genetic counseling in the Middle East.

## 1. INTRODUCTION

Genetic counseling is a specialised healthcare profession which provides information and support to individuals and families who are at risk of, or affected by, genetic conditions. While this field has gained recognition and acceptance in many parts of the world, it is still relatively new in the Middle East. In recent years, however, the demand for genetic counseling services has been growing in the region due to increasing availability of advanced genetic testing technologies to cope with the high prevalence of genetic diseases attributed to the practice of consanguinity and endogamy(El Naofal et al., 2023).

Typically, genetic counseling is provided by genetic counselors, who have specialized training and expertise in the field. However, in the Middle East, there are a limited number of genetic counselors available(Abou Tayoun et al., 2021; Abou Tayoun & Rehm, 2020). According to available statistics, there are currently around 40 genetic counselors practicing in Arab countries within the region supporting a population of over 450 million Arabs. Most of those counsellors practice in Saudi Arabia (N = 20)(Abacan et al., 2018; Abou Tayoun et al., 2021) and the United Arab Emirates, UAE (N = 10). There are currently no genetic counseling programs available in most Arab countries and genetic counselors practicing in this region have received training from programs around the world.

The extremely limited of availability of genetic counselors in the Middle East, already far below the global standard, is particularly stark given the outsized contribution of genetic diseases in this part of the world. To keep up with the demand, genetic counseling in the region is provided by physicians across various specialties. Little is known about the experience of healthcare professionals who provide these services. This lack of knowledge and understanding can hinder the improvement of genetic counseling services in the Middle East. Therefore, there is a pressing need to explore the perspectives of healthcare rofessionals who provide genetic counseling services to better understand the challenges they face and identify opportunities for improvement.

## 2. METHODS

### 2.1 Participants

An online questionnaire was created using google forms. This questionnaire (**Tables 1** and **2**) was distributed through email to healthcare providers ordering genetic testing across the Middle East. The responses were collected over a period of 5 months.

### 2.2 The Survey

The survey had two sets of questions. The first set focused on understanding the demographics of the responding provider and their patient population (**Table 1**). This included understanding provider backgrounds, patient and provider geographic information and the ethnicity and consanguinity levels of the patient population.

**Table 1:** Survey questions focusing on providers’ and patients’ demographics.

| Topic | Questions |
| --- | --- |
| Healthcare provider backgrounds | 1. What is your profession?<br>2. What setting do you practice in?<br>3. What speciality do you work in?<br>4. How long have you been practicing for?<br>5. How long have you been practicing for in the middle east? |
| Geographic information | 6. What is your nationality?<br>7. Which middle eastern country are you practising in?<br>8. Which country did you receive your training in?<br>9. Which countries did you previously practice in? |
| Ethnicity and consanguinity of patient population | 10. What ethnicities (e.g. Middle East, Africa, South Asia, South-East Asia, EasternEuropean, European, North American, South American do your patients belong to?<br>11. What is the estimated proportion of each ethnicity in your patient population?<br>12. What percentage of your patient population is consanguineous (e.g. parents are related by blood)? |

The second set of questions were the main survey questions (**Table 2**) and focused on understanding the cultural challenges in genetic counseling, patient’s attitudes towards genetics, any special counseling techniques, considerations employed by the providers, and the health care providers’ overall experience practicing in the Middle East.

**Table 2:**
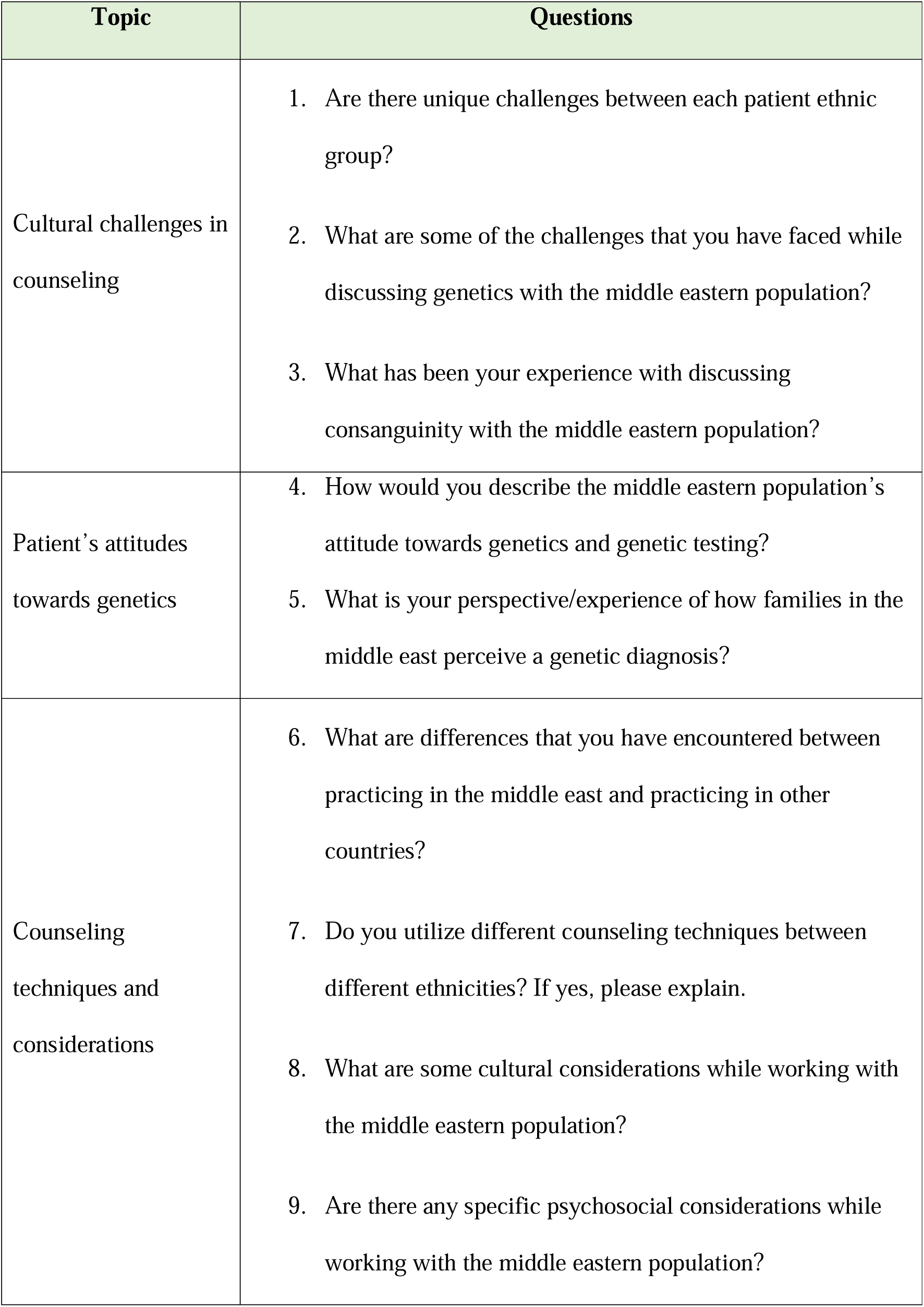
Survey questions focusing on genetic counselling practice.

| Topic | Questions |
| --- | --- |
| Cultural challenges in counseling | 1. Are there unique challenges between each patient ethnic group?<br>2. What are some of the challenges that you have faced while discussing genetics with the middle eastern population?<br>3. What has been your experience with discussing consanguinity with the middle eastern population? |
| Patient's attitudes towards genetics | 4. How would you describe the middle eastern population's attitude towards genetics and genetic testing?<br>5. What is your perspective/experience of how families in the middle east perceive a genetic diagnosis? |
| Counseling techniques and considerations | 6. What are differences that you have encountered between practicing in the middle east and practicing in other countries?<br>7. Do you utilize different counseling techniques between different ethnicities? If yes, please explain.<br>8. What are some cultural considerations while working with the middle eastern population?<br>9. Are there any specific psychosocial considerations while working with the middle eastern population? |

### 2.3 Data Analysis

All the answers were compiled and de-identified. Data were analysed using thematic analysis(Braun & Clarke, 2006). Answers were manually coded and organized by the first author (SS) and a code book was created. Answers were then sorted based on their content into categories. Themes were then derived across the categories.

## 3. SURVEY RESULTS

### 3.1 Participants

A total of 32 responses were received.

#### 3.1.1 Health care provider backgrounds

7 genetic counselors, 7 clinical genetics, 2 molecular geneticists, and 16 physicians across multiple specialties responded to the survey. Based on the results, half of the respondents identified themselves as being non-genetics healthcare professionals, 21.9% identified as clinical geneticists and another 21.9% were genetic counselors (**Figure 1**).

**Figure 1:**
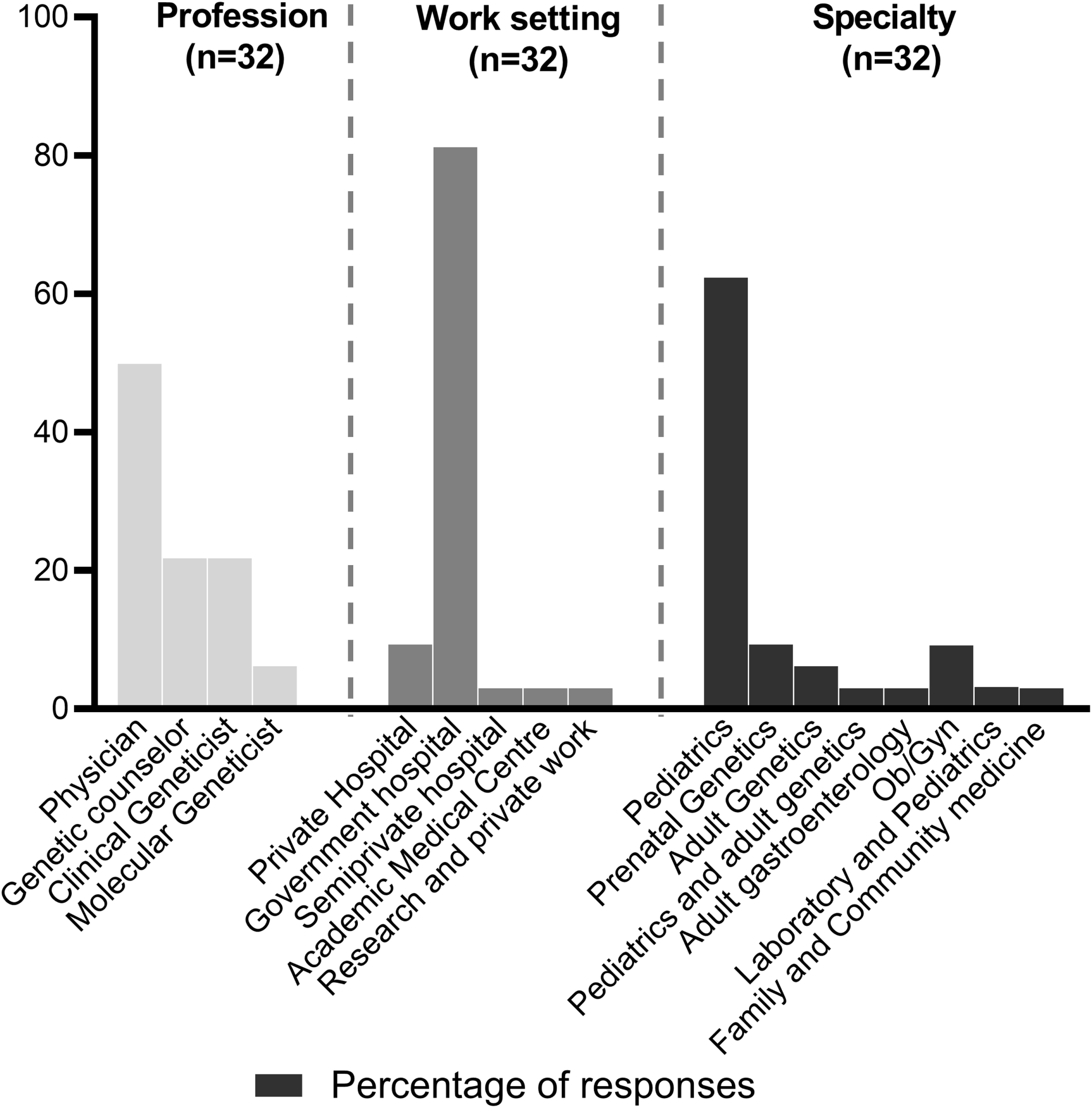
Number of respondents stratified by profession, work setting and specialty This bar graph illustrates the distribution of survey respondents across different professions, work settings, and specialties.

Most of the healthcare providers (81.3%) practised in government hospitals, while 9.4% providers practiced in private hospitals, 3.1% in a semi-private hospital, 3.1% in an academic medical centre and 3.1% in a research and private setting (**Figure 1**).

A majority of the providers (62.5%) worked in paediatrics followed by 18.7% in prenatal, obstetrics and gynaecology (**Figure 1**).

The responses regarding the duration of practice indicate that 37.5% of healthcare professionals participating in the survey have been practicing for 0-5 years, 25% for 6-10 years, 6.25% for 11-15 years, and 31.25% for more than 15 years. When considering practice specifically in the Middle East, 59.38% of respondents have been practicing for 0-5 years, 18.75% for 6-10 years, 3.13% for 11-15 years, and 18.75% for more than 15 years. (**Table 3**).

**Table 3.** Respondents’ years of experience.

| **Time frame** | **How long have you been practising for?**<br>**(% of respondents)** | **How long have you been practising in the Middle East?**<br>**(% of respondents)** |
| --- | --- | --- |
| 0-5 years | 37.5% | 59.38% |
| 6-10 years | 25% | 18.75% |
| 11-15 years | 6.25% | 3.13% |
| More than 15 years | 31.25% | 18.75% |

#### 3.1.2 Geographic information

Healthcare providers responding to this survey were from various nationalities currently practicing in the Middle East. Of the 32 respondents, 14 (43.75%) were from Saudi Arabia, followed by 4 (11.4%) from both the UAE and India, and 3 (8.6%) from Canada. The remaining respondents were from Ireland, the UK, Sudan, Turkey, and Palestine, with each country represented by one or two respondents (**Figure 2**).

**Figure 2:**
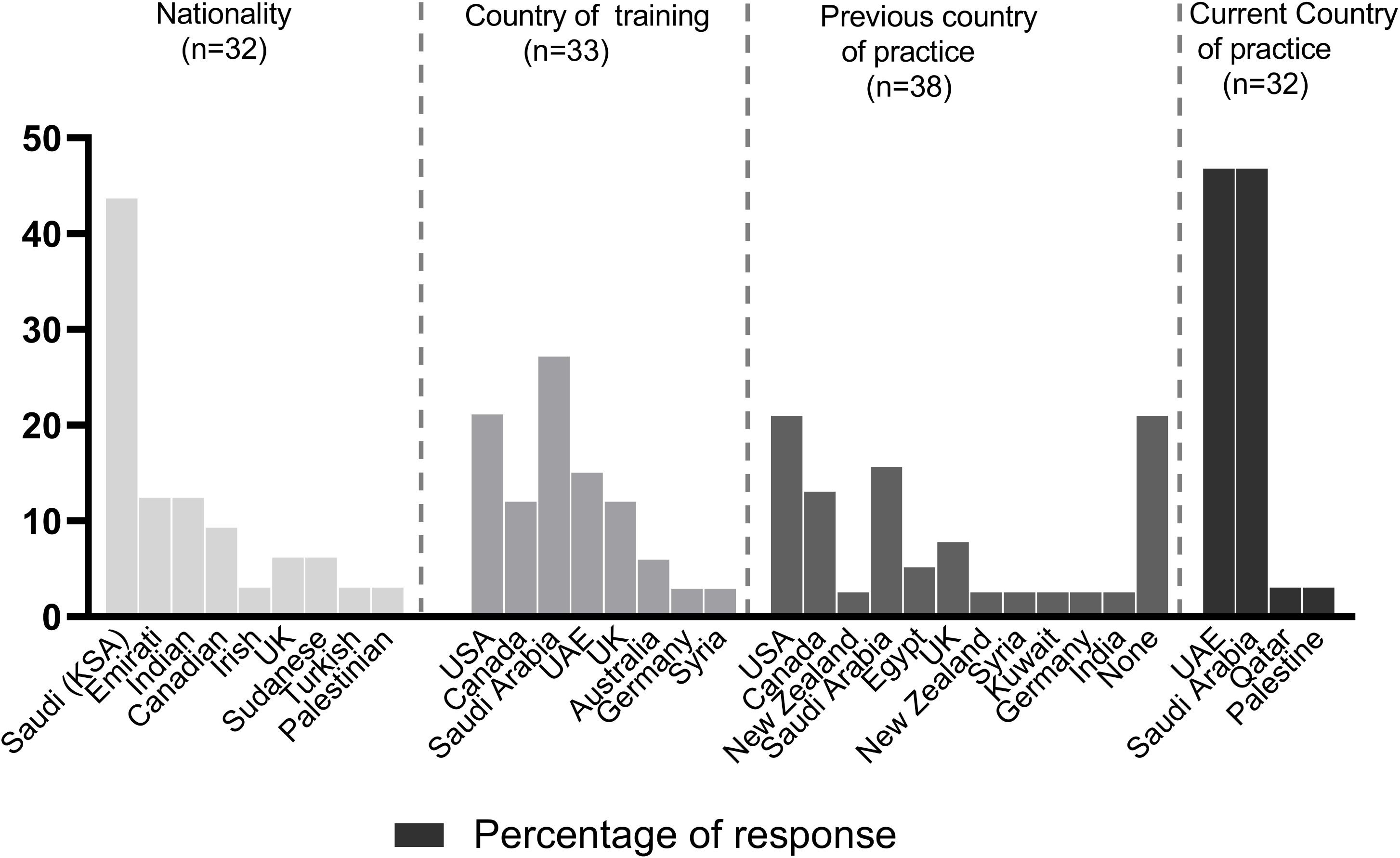
Number of respondents stratified by nationality, country of practice and training This bar graph showcases the distribution of survey respondents based on their nationality, country of training, previous country of practice, and current country of practice. Some respondents received training and practiced in multiple countries, and each country was considered as a separate entity in the analysis.

There are 15 respondents currently practising in the UAE, 15 in Saudi Arabia, 1 in Qatar, and 1 in Palestine (**Figure 2**).

Majority of healthcare providers received their training in the United States or Canada, accounting for 11 responses. Saudi Arabia is the next most common country where training was received, followed by the United Arab Emirates, United Kingdom, Australia, Germany and Syria. It is important to note that some respondents received their training in multiple countries (**Figure 2**). It is important to note that some respondents received training in multiple countries and each country was considered as a separate entity in the analysis.

Majority of the providers have previously practiced in either the USA or Canada (N = 13). Saudi Arabia was the third most common country where providers have previously practiced, with 6 responses. Egypt, UK, New Zealand, Syria, Kuwait, Germany, and India were also mentioned as previous practice locations. Additionally, 8 providers responded with “none”, indicating that they have not practised in any other country previously (**Figure 2**). It is important to note that some respondents practiced in multiple countries and each country was considered as a separate entity in the analysis.

#### 3.1.3 Ethnicity and consanguinity of patient population

Based on the responses provided by providers currently practising in Saudi Arabia, the majority of the patients served by these providers belong to Arab ethnicities, with percentages ranging from 90% to 100%. Some respondents also mentioned serving a small proportion of patients from other ethnic backgrounds, such as Indian, Pakistani, European, American, and African. A few respondents did not provide specific numbers or details regarding the ethnicities of their patients. One respondent noted that the patients served are predominantly Muslim, with a small percentage of Christians.

Based on the responses provided by healthcare providers in the UAE, their patients are from diverse ethnic backgrounds. The majority of patients are reported as Arab, with percentages ranging from 70% to 100%. South Asian patients were also commonly reported, ranging from 10% to 20%. Patient ethnicities, other than Arabs, mentioned included Caucasian, Pacific Islander, Filipino, European, Indian, Pakistani, African, and Southeast Asian. Some healthcare providers reported that their patients were multicultural or multinational.

According to a provider from Qatar, the patient population in this country comprises of multiple ethnicities, but the majority are of Middle Eastern/North African origin.

Approximately 70% of the patients are estimated to be of Middle Eastern/North African origin, while the remaining 30% belong to other nationalities, with East and Southeast Asians being the majority and Europeans being the minority (less than 5%).

The provider from Palestine reported that their patients are predominantly Arab Palestinians but did not provide a specific percentage.

The survey question “What percentage of your patient population is consanguineous?” received a range of responses, with some healthcare providers estimating that almost all of their patient population was consanguineous, while others reported percentages as low as 10%. The majority of responses indicated a high percentage, with many estimating around 50-60%, 80%, or even more than 95%. Some providers noted that they did not have statistics available, but estimated percentages ranging from 25% to over 75%.

### 3.2 Provider’s views

#### 3.2.1 Attitudes towards genetic testing

Out of the total responses analysed in the survey, 19 of them (59.4%) indicated that patients have a positive attitude towards genetic testing.

These responses expressed a willingness and interest in pursuing genetic testing, regardless of factors such as ethnicity or insurance coverage. Some respondents believed that most patients are interested in getting tested, while others reported an increase in awareness and acceptance of genetic testing among their patient populations. Some respondents noted that individuals and couples are open to genetic testing, with some families willing to pursue the most comprehensive testing available. Some respondents also reported that there is still limited understanding regarding genetic testing among patients.

However, there are also negative attitudes towards genetic testing. As part of the thematic analysis, it was found that six individuals mentioned stigma associated with a genetic diagnosis. These individuals pointed out that there were cultural and social factors that influenced patients’ willingness to pursue genetic testing. For example, some traditional families had a stigma against genetic testing and were not keen on pursuing carrier testing. In some paediatric cases, families may be reluctant to undergo testing, especially if it involves parental samples. In addition, families were less open about medical issues in the family history due to possible stigma associated with a diagnosis. Furthermore, there were concerns regarding the impact of genetic ’blame’, and concerns regarding marital prospects for girls.

#### 3.2.2 Cultural considerations

##### 1. Consanguinity

One recurring theme in the responses was the prevalence of consanguinity within the Middle Eastern patient population. While families often understand the potential risks associated with consanguinity, it appears that this knowledge is not always acted upon. Some respondents noted that highly educated individuals and the general population still hold the belief that consanguinity marriage is the best option.

Respondents also noted that while consanguinity cannot always be prevented, people are becoming more understanding of the risks. However, this awareness has not necessarily resulted in changes in behavior or attitudes towards consanguinity. It was also observed that consanguinity is a normalized and prevalent part of life within this region and is not stigmatized.

Some respondents felt that the Middle Eastern population is generally aware of and accepting towards the risks associated with consanguinity. However, there were also some challenges noted, such as patients unknowingly denying consanguinity if the consanguinity is a third-degree relation or higher. Providers may need to ask this question multiple times in different ways to obtain an accurate answer. Another challenge noted was when a family has a known genetic disorder, and members of the same family still wish to marry without confirming their carrier status with genetic testing. This scenario was described as not uncommon and could present a challenge for providers.

Other cultural considerations that were noted in the survey include polygamy and larger families.

##### 2. Polygamy

Polygamy is a practice where a man is allowed to marry more than one woman at a time, and it is still legal and practiced in some Arab countries. Polygamy was brought up by one responder who stated that in some cases, fathers may consider marrying another woman for various reasons, including economic or social status, and they may use genetic testing results as a justification to do so.

##### 3. Larger families

Larger families can provide extended family and social support networks for patients but can also make it difficult for parents to manage if they have a child with a genetic disorder.

Additionally, it can be harder for families to accept a genetic diagnosis if other children in the family are healthy or typical.

#### 3.2.3 Closed / Private

Seven responders to the survey highlighted a cultural consideration specific to the region, where individuals tend to be more private or closed. The feedback revealed three main themes.

The first theme indicates that people in this region are generally less willing to be open about their medical issues, including their family history. Arab populations can be hesitant to pursue parental testing or disclose medical information due to the possible stigma associated with a diagnosis. One respondent shared their personal experience, stating that they find it difficult to establish a connection with the Arab population and provide psychosocial counseling, as patients are often reluctant to share their coping mechanisms. Additionally, discussing planning for future pregnancies can be challenging, and some patients may not be comfortable having such conversations with their physician.

The second theme that emerged is related to the reluctance of individuals in this region to be open to “outsiders”. Some respondents noted that cultural barriers exist for individuals outside the Arab community, and patients may feel less willing to open up to them. This can make it challenging to establish rapport and provide effective psychosocial counseling.

However, some respondents did not face any barriers in this regard, as they were seen as part of the Arab community and communication was easy. Others mentioned the cultural trait of stoicism, which can make it difficult to engage with patients from the Middle Eastern population, as they may not be accustomed to discussing their emotions and feelings. As a result, some respondents felt like outsiders while interacting with this population.

The third theme that emerged from the responses is that individuals in this region can sometimes be less willing to open up to their family members. Despite strong family ties, there is often a reluctance to share information about a child’s genetic diagnosis or condition, which can make it challenging to provide appropriate counseling and care. Some individuals do share the information and take advantage of available preventive measures, while others may be too protective of the information, preventing other family members from benefiting from it.

#### 3.2.4 Language barrier

Based on the survey responses, 5 individuals brought up the issue of language barrier as a challenge in communicating with patients. They noted that this barrier can affect the quality of communication and understanding between healthcare providers and patients, especially for non-English speaking patients.

One respondent mentioned that a language barrier often leads to more direct communication, with less focus on psychosocial aspects of a session.

Another respondent noted that language is a barrier in their practice, as they do not have formal interpreters and have to rely on nurses for interpretation, which can lead to challenges such as the nurses adding or subtracting information on their own or having conversations with patients without interpreting. Additionally, the way information is explained may need to be adapted depending on the language barrier.

Others noted that low levels of genetic literacy and cultural differences can compound the issue of language barriers, resulting in additional challenges in genetic counseling.

#### 3.2.5 Coping with a genetic diagnosis

In the survey question asking about how individuals cope with a genetic diagnosis, 16 out of 32 (50%) responses indicated that families generally cope well. The reasons for this are varied and include existing family history, positive religious beliefs, and the severity of the disease. Many respondents noted that families who have had exposure to genetic diseases through close relatives are better equipped to cope with a diagnosis. This is particularly true for Arab families, who often readily accept the diagnosis and pursue all options for treatment. One genetic counselor compared their experience in the Middle East to their practice in North America where the majority of families who have no family history of genetic disease may be less prepared for a diagnosis.

Religious beliefs were also noted as a factor in coping with a genetic diagnosis. Many Arabic Muslim patients express the belief that their diagnosis was “God’s will” and that this belief played a significant role in helping them cope. Respondents also noted that families who dealt with the diagnosis from a primarily religious point of view tended to be more accepting of the situation and optimistic about finding a cure in the future.

According to the survey results, limited resources pose a significant challenge for individuals coping with a genetic diagnosis. Four survey respondents highlighted the lack of clinical genetics support, multidisciplinary clinics, rehabilitation centers, and support groups for genetic disorders in the Middle East as major issues. In addition, insufficient family support, particularly from husbands, was identified as a concern. Financial burden was also mentioned as a potential obstacle to coping effectively with a genetic diagnosis.

#### 3.2.6 Low genetic literacy

A total of 11 out of 32 (34.4%) responses expressed concerns about the low levels of genetic knowledge and understanding among patients and healthcare professionals in the region. Some respondents noted that patients have misconceptions about the possibility of treating genetic diseases and the concept of consanguinity. Others mentioned the lack of awareness and education around genetic services among both patients and providers. Additionally, some healthcare professionals noted that families in the region need more orientation and education about genetics and genetic testing.

## 4 ANALYSIS OF RESULTS AND DISCUSSION

### 4.1 Participants

The majority of the healthcare providers who participated in the survey (93.75%) are currently practicing in either the UAE or Saudi Arabia, with each country having an equal number of responses (15 each). It is important to note that the relatively small sample size of the survey limits the generalizability of these findings and further research is needed to gain a more comprehensive understanding of the distribution of healthcare providers throughout the Middle East.

The survey findings suggest that the field of genetics in the Middle East is diverse in terms of profession, setting, and specialty, with genetic counselors being just one component (21.9%) of a broader range of healthcare professionals working in genetics-related fields.

The findings suggest a mix of both early-career professionals and those with extensive experience in the field and a higher proportion of relatively new practitioners in the Middle East, with a significant influx of healthcare professionals with limited experience in the region.

Genetic counseling in the UAE is provided by a diverse and international workforce, with professionals trained in various countries and having experience practicing in different parts of the world. This diversity may bring a range of perspectives and expertise to the field of genetic counseling in the Middle East, which can ultimately benefit patients and families seeking genetic counseling services.

The surveyed healthcare professionals in the Middle East represent diverse nationalities, highlighting the multicultural nature of genetic counseling practices in the region. It is noteworthy that providers have received training from various countries across the globe, reflecting a global exchange of expertise and experiences.

It appears that the patient population in the UAE is diverse and reflective of the country’s status as a cosmopolitan hub. The responses suggest that the providers in Saudi Arabia primarily serve patients of Arab ethnicities, but also serve patients from a diverse range of backgrounds. Patients in Qatar are mainly of Middle Eastern/North African origin, while those in Palestine are predominantly Arab Palestinians. The responses suggest that consanguinity is a prevalent issue, with estimates ranging from 10% to almost all of the patient population.

### 4.2 Provider’s views

The study reveals a generally positive attitude towards genetic testing among patients and families, regardless of ethnicity or insurance coverage, with many expressing interests in pursuing it. Negative attitudes towards genetic testing are also observed, influenced by stigma and social concerns associated with inherited diseases, as well as language and cultural barriers. Addressing negative attitudes and barriers to testing is crucial for healthcare providers to ensure access to information and support patients in making informed decisions about their health.

Consanguinity is a prevalent and normalized practice in the Middle East, posing challenges for genetic counseling due to families’ reluctance to act on knowledge of associated risks. Genetic counselors need culturally sensitive approaches to educate families about the risks of consanguinity and encourage genetic testing before marriage. Raising awareness about the risks and available testing options can empower families to make informed decisions and reduce the impact of consanguinity-related genetic disorders.

When discussing polygamy in the Arab population, cultural sensitivity is crucial to address the complexity of the issue and provide accurate information about genetic testing. In the Middle Eastern patient population, larger families present challenges for families with genetic disorders, and genetic counselors may face difficulties in coordinating testing for extended family members due to the presence of larger families.

Overall, these cultural considerations highlight the need for sensitivity and cultural awareness in genetic counseling. It is important to approach each patient and family with an understanding of their unique cultural background and to work collaboratively with them to provide the best possible care.

The survey responses indicated another cultural consideration that individuals in the region may tend to be more private or closed. This was reflected in three themes. The first theme is that people in the region are less willing to be open about medical issues in the family history, particularly among Arab populations, which can be attributed to possible stigma associated with a diagnosis. The second theme highlights that people in the region can be less willing to open up to non-Arab outsiders. The third theme suggests that people in the region can be less willing to open up to their family members, particularly when their child is the first to be diagnosed with a genetic condition. This can lead to challenges in communication and genetic counseling. It may be difficult to establish a rapport and build trust with the patient, which is essential for effective counseling. Patients may be less willing to share personal information, such as their family history, which can hinder the identification of genetic risk factors. Additionally, cultural beliefs and stigma surrounding genetic disorders may prevent patients from seeking counseling or following through with recommended actions. Finally, language barriers and limited access to genetic testing services may further compound these challenges. Therefore, healthcare providers working with Arab patients must be culturally sensitive and adept at navigating these complexities to provide effective counseling services.

Language barrier was a commonly mentioned issue among the respondents. Language barrier can have an impact on rapport building, information transfer, and psychosocial counseling.

The combination of language barriers, low levels of genetic literacy, and cultural differences can create distinct difficulties in genetic counseling. Overall, these responses highlight the importance of addressing language barriers and finding effective ways to communicate about genetic testing with diverse patient populations.

The survey results indicate that families in the Middle East generally cope well with a genetic diagnosis, with factors such as existing family history, positive religious beliefs, and severity of the disease influencing their coping mechanisms. Arab families, in particular, tend to accept the diagnosis and actively seek treatment options. However, limited resources, including lack of clinical genetics support, rehabilitation centres, and support groups, pose significant challenges for individuals coping with a genetic diagnosis in the region. Financial burden and insufficient family support were also identified as concerns.

The survey responses suggest that there is a lack of genetic awareness and literacy among patients and healthcare professionals in the Middle East. This could potentially lead to lower compliance rates and difficulty in obtaining accurate family history information, which is crucial for genetic counseling. The survey responses highlight the need for more efforts to increase genetic awareness and literacy in the Middle East. To address these concerns, further research is needed to understand the specific barriers to genetic education and awareness in the Middle East, as well as the most effective strategies for increasing genetic literacy and improving access to genetic counseling services.

## 5. CONCLUSION

In summary, our survey revealed that the majority of healthcare providers offering genetic counseling services in the Middle East come from diverse training backgrounds, and while they primarily counsel Middle Eastern Arab patients, they also work with other ethnicities in specific countries. Overall, healthcare providers reported that patients in this region have positive and accepting attitudes towards genetic testing. They demonstrated awareness and adaptation to social attributes such as polygamy and consanguinity, which are prevalent in the population. However, language barriers and social stigma emerged as significant obstacles that require further efforts to overcome in order to maximize the benefits of genetic testing and counseling in this population.

The field of genetic testing is continuously evolving, emphasizing the need for genetic counseling services to adapt and expand accordingly. Strategies like standardizing guidelines, increasing awareness, and investing in education can address the challenges of genetic counseling in the Middle East. However, further research is necessary to comprehensively understand the cultural considerations and develop tailored interventions that improve access to genetic services and enhance awareness and understanding of genetic conditions in the region.

## 6. LIMITATIONS

The limitations of the survey include the small sample size of only 32 responses, which may not be representative of all healthcare professionals in the Middle East. Additionally, the survey was limited to only four countries (United Arab Emirates, Saudi Arabia, Qatar, and Palestine) with only 1 response each from the latter two countries, and therefore may not reflect the attitudes and experiences of healthcare professionals in other Middle Eastern countries. Furthermore, the survey was focused on healthcare professionals’ perspectives rather than those of patients and their families, which may have provided a more comprehensive understanding of the challenges and considerations surrounding genetic counseling in the region. Lastly, self-report bias may have influenced the responses, as participants may have provided socially desirable answers or may not have accurately represented their experiences and attitudes towards genetic counseling.

## AUTHOR CONTRIBUTIONS

Authors Shruti Shenbagam and Ahmad N. Abou Tayoun confirm that they had full access to all the data in the study and take responsibility for the integrity of the data and the accuracy of the data analysis. All the authors gave final approval of this version to be published and agree to be accountable for all aspects of the work in ensuring that questions related to the accuracy or integrity of any part of the work are appropriately investigated and resolved.

## ACKNOLEDGEMENTS

We would like to express our sincere gratitude to all the healthcare professionals who took part in this study and generously shared their insights and experiences. Without their contributions, this research would not have been possible. We are grateful for their time and willingness to provide valuable information.

## HUMAN STUDIES AND INFORMED CONSENT

This study was reviewed and granted an exemption by the Institutional Review Board at Mohamed Bin Rashid University (IRB-2023-152). All procedures followed were in accordance with the ethical standards of the responsible committee on human experimentation (institutional and national) and with the Helsinki Declaration of 1975, as revised in 2000. Implied informed consent was obtained for individuals who voluntarily completed the online survey and submitted their responses.

## DATA AVAILABILITY STATEMENT

The data that support the findings of this study are available on request from the corresponding author. The data are not publicly available due to privacy or ethical restrictions.

